# Antimicrobial consumption in pediatric intensive care units during the first year of COVID-19 pandemic

**DOI:** 10.1101/2021.10.11.21261865

**Authors:** MFM Porto, CSM de Oliveira, MEO Pires, RRQ Santos, CH Teixeira, CV Souza, AR Araujo da Silva

## Abstract

**Introduction:** The absence of standardized treatment for critical children admitted in pediatric intensive care units (PICUs) with COVID could lead to an increase in antimicrobial consumption, as indirect effect.

**Aim:** To describe trends of antimicrobial consumption in two PICUs before and during the COVID pandemic year.

**Methods:** We did a retrospective study in children admitted in two PICUs of Rio de Janeiro city, between March 2019 and March 2021. The first year represented the pre-pandemic period and the last one the pandemic period. Trends of antimicrobial consumption were measured by days of therapy (DOT/1000 patient-days) and analyzed by linear regression for antibiotics, antivirals and antifungals

**Results:** Number of patients-days in the PICU 1 was 3495 in the pre-pandemic period and 3600 in the pandemic period. The overall DOT/1000 PD of antibiotics, antivirals and antifungal was 15,308.1, 942.8 and 1,691.1, respectively in the pre-pandemic period and 13,481.5, 1,335.4 and 1,243.7, respectively in pandemic period. It was verified trend of reduction of antibiotic and antifungals and increase in antivirals consumption. Number of patients-days in the PICU 2 was 5029 in the pre-pandemic period and 4557 in the pandemic period and the overall DOT/1000 PD of antibiotics, antivirals and antifungal was 16,668.5, 1,385 and 1,966.7, respectively in the pre-pandemic period and 10,896.5, 830.7 and 677.3 in pandemic period. It was verified trend of reduction of antibiotic, antivirals and antifungals consumption.

**Conclusion:** Trends of antimicrobial consumption reduction were verified for antibiotics and antifungals in two PICUs and reduction for antiviral in one of them

## Introduction

Since the first description of systematizated interventions destinated to improve antimicrobial prescriptions in hospitals, using a package of measures nominated as antimicrobial stewardship programms (ASPs), several similar initiatives were developed around different healthcare institutions of the world. ^1,2,3^

The ASPs were also implemented in children hospitals with decrease of broad spectrum and/or target antimicrobials consumption. Hersh et al compared antibiotic prescribing rates in a group of pediatric hospitals with formalized ASPs and without formalized ASPs, reporting declines in antibiotic use in 8 of 9 hospitals with ASPs, with an average monthly decline in days of therapy/1000 patient-days of 5.7%. ^4^ Chautrakarn and cols reported a significant decrease of vancomycin, colistin and meropenem use after a 6-month period of ASP implementation, in a tertiary pediatric hospital in Thailand. ^5^ 2019).

Children are less affected than adults in the COVID pandemics, but unusual presentation and critical conditions as multisystem inflammatory syndrome (MIS-C) could be present, causing admission in pediatric critical care units. ^6^ Considering that initial presentation of MIS-C could mymics other critical conditions as sepse and pneumonia, and no specific treatment is available for COVID in chldren, it’s possible that antibiotic use may have increased during the first year of pandemic. ^7^

The aim of this manuscript is to describe trends of antimicrobial consumption in two pediatric intensive care units (PICUs) before and during the COVID pandemic year, in two healthcare institutions with a previous implemented ASP.

## Material and methods

### Study design and setting

We conducted a retrospective cohort study in two pediatric hospitals in Rio de Janeiro, city, Brazil, between March 1, 2019 and March 31, 2021.

Institutions and intensive care units classification:

1. Prontobaby Hospital da Criança (Rio de Janeiro, Brazil) is a 135-bed private hospital including one PICU (10-bed capacity, named as PICU 1). The unit receives clinical and surgical patients referred from its own wards and from other services. This hospital hosts medical students from Universidade Federal Fluminense for practical classes.
2. Centro Pediátrico da Lagoa (Rio de Janeiro, Brazil) is a 39-bed private hospital including a 15-bed PICU (named as PICU 2), with the same profile as Prontobaby PICU patients.

The inclusion criteria were PICU admission and antimicrobial use for more than 24 hours. Children that used topic and inhaled antibiotics were excluded. Antimicrobials were analysed separately as antibiotics, antivirals and antifungals

### Definition of metrics

We calculated DOT for each antibiotic used (oral, or intravenous) and DOT/1000 PD in the intensive care units during the stay of children in each unit, according the Polk et al definition. ^8^

### Antimicrobial stewardship programm

A systematized antimicrobial stewardship program is in progress in both hospitals, since November 2017. The main components of the ASP are: written guidelines for antimicrobial use, rounds and discussion of cases, antibiotic restriction policy, de-escalation of antibiotic, support of microbiology laboratory and training of staff.

### Antibiotic policy restriction

The following antimicrobials were included in an antimicrobial restriction policy, requiring pre-approval by a paediatric infectious disease specialist: Amphotericin B lipid formulations, Caspofungin, Ceftazidime/Avibactam, Ceftobiprol, Colistin (inhaled), Daptomycin, Ertapenem, Imipenem, Linezolid, Meropenem, Micafungin, Polymyxin B, Teicoplanin, Tigecyclin, Voriconazole.

### Data source and statistical analysis

A descriptive analysis was performed using Microsoft Excel. When appropriate, we used Chi square test for categorical variables and Mann–Whitney U test for continuous variables. Trends of consumption were analysed by linear regression.

### Ethical approval

The study was submitted and approved by Ethics Committee of Faculty of Medicine (Universidade Federal Fluminense) and Prontobaby Group, under number 5.022.885 dated from October 6, 2021. Data of antimicrobial consumption were obtained from the electronic system of the two hospitals anonymized, without patient information

## Results

Between the period study, 2,547 patients were admitted in both PICUs, representing 16,681 patient-days. The table 1 shows number of admitted patients and patient-days per year and per unit.

**Table 1.** Number of admitted patients and patient-days, per year in two PICUs fo Rio de Janeiro (March 2019-March 2021)

|  | PICU 1 |  |  | PICU 2 |  |  |
| --- | --- | --- | --- | --- | --- | --- |
|  | Pre-pandemic<br>period | Pandemic<br>period | Total | Pre-pandemic<br>period | Pandemic<br>period | Total |
| Patients | 468 | 617 | 1,085 | 724 | 738 | 1,462 |
| Patient-days | 3,495 | 3,600 | 7,095 | 5,029 | 4,557 | 9,586 |

In the figure 1, we show trends of consumption of antibiotics, antivirals and antifungal in PICU 1 and in the figure 2 the pattern of consumption in PICU 2.

**Figure 1.**
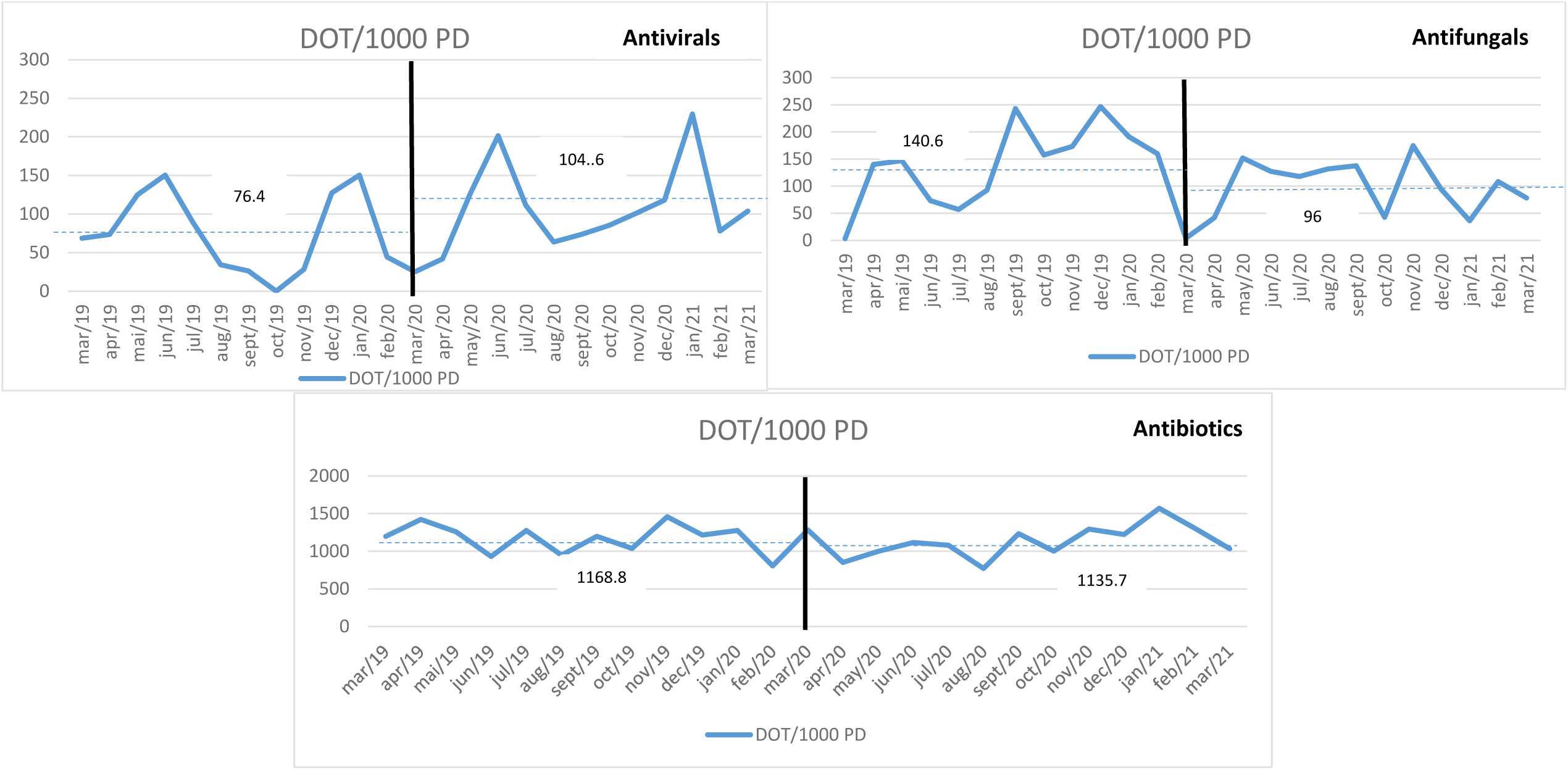
Trends of antimicrobial consumption in the PICU 1 (March 2019-March 2021) Vertical black line: Beginning of pandemics in Brazil, Blue traced line: Mean of consumption (in DOT/1000 patient-days)

**Figure 2.**
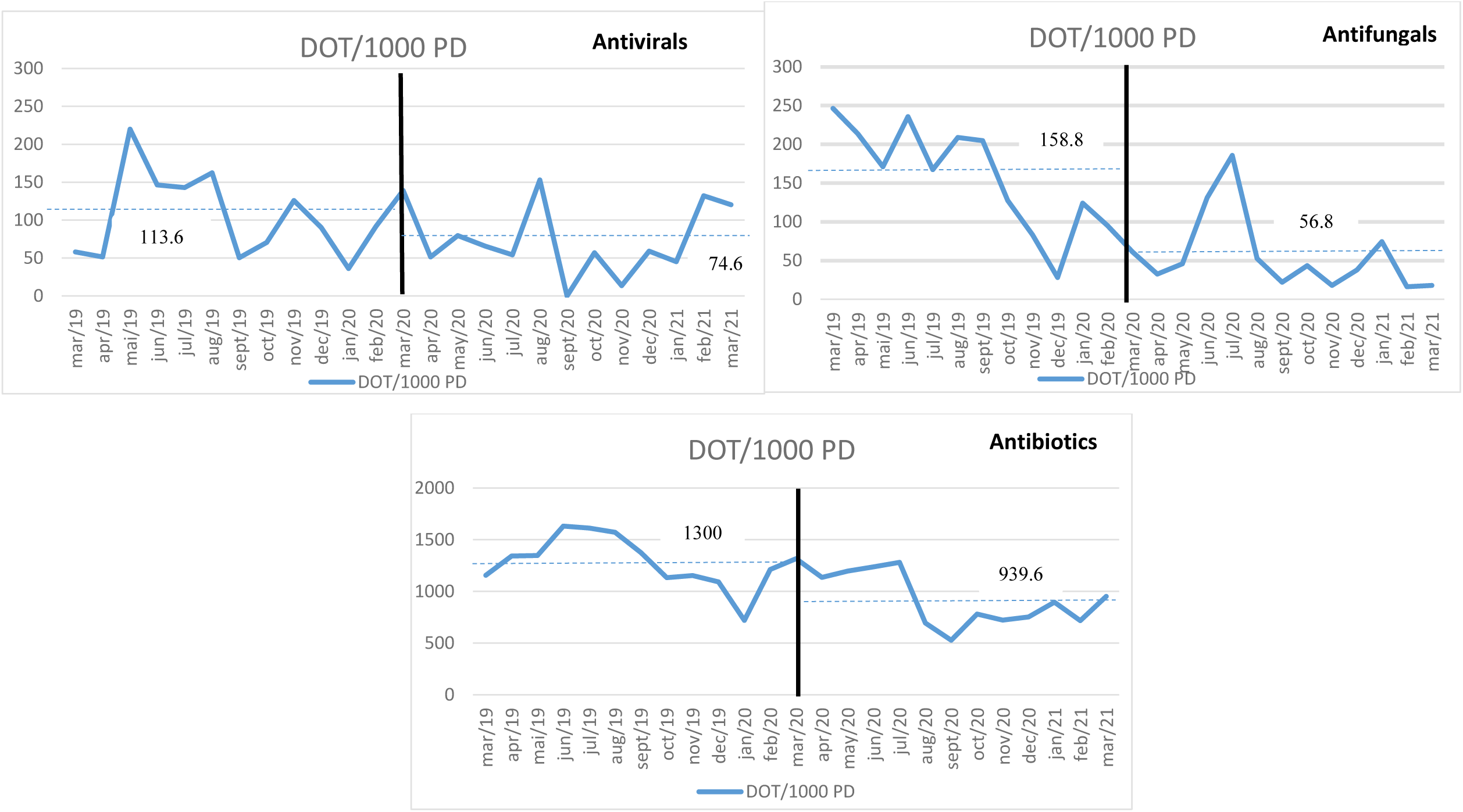
Trends of antimicrobial consumption in the PICU 2 (March 2019-March 2021) Vertical black line: Beginning of pandemics in Brazil, Blue traced line: Mean of consumption (in DOT/1000 patient-days)

## Discussion

In this paper, we explore the influence of COVID-19 pandemics on antimicrobial consumption of three main classes: antibiotics, antivirals and antifungals in two PICUs with previous antimicrobial stewardship. Considering that respiratory symptoms are present in most pediatric COVID-19 cases admitted in PICUs and, in many cases, these same symptoms are undistinguishable from other syndromes as bacterial pneumonia, use of antimicrobials could increase in the pandemic period.

The increase of antibiotic use during the first year of pandemic was verified in a recent systematic review conducted by Giacomelli and cols where the analysis of 28,093 patients from November 2019 to December 2020 found that 58.7% received antibiotics, ranging from 1.3% to 100% coverage. ^9^ Castro-Lopes et al, studying antibiotic consumption in a single center of Portugal, verified statistically significant increases in 2020 for total antibacterials, macrolides, cephalosporins, amoxicillin/clavulanic acid, carbapenems, meropenem, and third-generation cephalosporins, while a reduction was seen in cefazolin/cefoxitin. ^10^

Our ASP was implemented in 2017 and since then, a rigorous surveillance of antibiotics prescribing is in progress, involve a multidisciplinar team (physicians, nurses and pharmacists), and includes patients admitted in pediatric intensive care. Despite the relevance of ASPs, few studies are available in children specially to verify reduction in antibiotic consumption and use of broad-spectrum/restricted agents in PICUs. ^11^

We verified trends of antibiotic reduction in out two PICUs, being more notable in PICU 2. In contrast to our results, Verlasco-Arnaiz and cols described increase in antibiotic consumption between February to April 2020 in a PICU of Spain, in comparision of the same period in 2019 (pre-pandemic year). ^12^

Trends of antifungals consumption were also noted in both PICUs studied. Despite antifungal consumption in PICUs is less reported than antibiotic consumption, there a recente concern about emergence of Candida auris resistant to different antifungals, also reported in pediatric and neonatal settings. ^13,14^

When antiviral consumption were analyzed, we verified a trend of reduction in one PICU and increase in the second one. The reason of this finding could be attributable due to empirical treatment of children with respiratory acute distress syndrome with Oseltamivir and local difficulty to identify Influenza vírus in respiratory secretion. Consumption of Oseltamivir reached 43% of children admitted in 19 Brazilian PICUs with MIS-C. ^6^ In other countries as USA, where Remdesevir is available, similar percentage of antiviral use (46.2%) was reported in children admitted in PICU. ^15^

Limitation of this article includes that the findings reflect local reality of two major PICUs of Rio de Janeiro city. Another limitation was having carried out the study in hospitals with a well established ASP and stricted control of antimicrobials, which not to be the reality of many hospitals.

We conclude that even during the first year of COVID pandemics, trends of antimicrobial consumption reduction were verified for antibiotics and antifungals in two PICUs and reduction for antiviral in one of them.

## Data Availability

All data are available, if necessary

## Acknowlegdment

We thank to Mario Eduardo Viana for supporting this research.

## Notes

### Competing Interest Statement

The authors have declared no competing interest.

### Funding Statement

No funding was obtained for the current research

